# Sleep Quality and Psychological Distress in Chinese Nursing Interns: The Moderating Effect of Social Support in the Association with Anxiety and Depression

**DOI:** 10.64898/2026.03.06.26347775

**Authors:** Yanyan Zhao, Fuzhi Liu, Lanlan Chen, Xiuli Li, Zhuote Tu, Biyu Wu

**Author notes:** Correspondence: Biyu Wu. These authors contributed equally to this work (Yanyan Zhao, Fuzhi Liu).

## Abstract

**Background:** Nursing interns are at high risk of psychological distress due to academic and clinical stressors. While poor sleep quality is linked to anxiety and depression, the buffering role of social support remains underexplored in this population.

**Aims:** To explore the role of social support in regulating the relationship between sleep and mental health among nursing interns.

**Methods:** A total of 396 nursing interns completed self-administered questionnaires including the Pittsburgh Sleep Quality Index (PSQI), Social Support Rate Scale (SSRS), Generalized Anxiety Disorder-7 (GAD-7), and Patient Health Questionnaire-9 (PHQ-9). Hierarchical regression and simple slope analyses were used to test moderation effects.

**Results:** Poor sleep quality was significantly associated with higher anxiety (β=0.449, *P*<0.001) and depression (β=0.535, *P*<0.001). Social support significantly moderated these relationships. Under low social support, the effects of sleep quality on anxiety (β = 0.602) and depression (β = 0.779) were stronger than under high support (anxiety: β = 0.396; depression: β = 0.515).

**Conclusions:** Social support buffers the adverse psychological effects of poor sleep among nursing interns. Interventions should integrate sleep hygiene education with strategies to enhance social support.

## 1. Introduction

In the field of medical care, nursing interns are the backbone of the future nursing team, and their physical and mental health status is directly related to the quality and efficiency of nursing services^[1]^. In recent years, with the continuous change of nursing education mode and the increasing internship task, nursing interns are facing unprecedented pressure and challenges. Sleep quality is not only a key indicator of physical and mental health, but also an important determinant of mental health, which is particularly prominent among health care workers under high levels of occupational stress. Existing research has clearly confirmed that sleep quality is not only a crucial determinant of mental health but also a key support for psychological resilience in professional groups, especially for healthcare workers who are often under prolonged high-pressure conditions. Sleep quality often serves as a “barometer” of occupational mental health^[2-3]^. A cross-sectional survey showed that poor sleep quality was significantly positively correlated with anxiety (OR=1.38) and depression (OR=1.58, P=0.001)^[4]^. A multi-center survey covering over 7,000 individuals in China revealed that 65.8% of clinical nurses experience sleep problems. Network model analysis suggests prioritizing interventions for insomnia and sleep disorders can help alleviate anxiety and depressive comorbidities^[5]^. In addition, a large sample of longitudinal cohort studies found that poor sleep quality decreased from 18.3% to 16.2% (P <0.05). Depression symptoms and anxiety symptoms also improved (P <0.05)^[6]^. Nursing interns are in a vulnerable stage of transition from academic training to clinical practice, facing multiple stressors such as work overload, role ambiguity, emotional labor and evaluation pressure. Their sleep-psychological path is more specific, and their sleep rhythm is easily disturbed, leading to sleep disorders^[7]^. Many domestic and foreign research data show that the incidence of poor sleep quality among nursing interns is high^[8-10]^. In a four-year longitudinal study, it was found that the sleep quality of nurses showed a continuous decline, especially during the transition from clinical internship to work-life, and the decline was most obvious^[11]^. Compared with nurses, due to the special clinical learning environment, nursing interns have shorter sleep time, significantly reduced sleep efficiency and more sleep interruption^[12-13]^. Therefore, sleep quality is a key predictor of mental health in healthcare workers, especially nursing interns who are more prone to sleep disorders due to multiple stressors. It is necessary to prioritize sleep interventions to reduce the risk of anxiety and depression.

Social support refers to the emotional, psychological or physical support provided by individuals or organizations, which has a certain impact on individual mental health^[14]^. The social support for nursing interns encompasses multiple dimensions, which can be categorized into structural attributes (social integration and support networks) and functional attributes (educational support, psychosocial support, and instrumental support). Its effectiveness depends on prerequisites such as stress and crisis, personal needs, social networks, and social climate, while yielding positive outcomes that improve mental health and enhance quality of life^[15]^. As a key group in the medical system, nursing interns often suffer from great psychological burden due to the intensity of clinical training, frequent shifts and heavy academic pressure. These pressures are easy to lead to poor sleep quality, which will aggravate psychological problems such as anxiety and depression^[16-17]^. Although some studies have shown that social support can improve sleep quality and reduce anxiety and depression levels^[18-21]^. Nevertheless, the precise mechanism through which sleep quality interacts with psychological issues, including anxiety and depression, and specifically its potential regulatory role, merits further study. Existing evidence primarily originates from other populations^[22-24]^, while systematic research on the moderating effect of social support in the “sleep-mental health” pathway among nursing interns remains limited. Crucially, determining whether social support can mitigate psychological impacts caused by sleep disorders is pivotal for developing targeted interventions to enhance mental well-being within this population—a gap that current studies have yet to address adequately.

This study focuses on the group of nursing interns in China, aiming to explore the interaction between sleep quality, social support, anxiety, and depression, with particular attention to the moderating role of social support in the relationship between sleep quality and mental health outcomes (anxiety and depression). We hypothesize that social support plays a significant moderating role. When social support is low, poor sleep quality shows stronger associations with psychological distress, while high social support weakens this correlation. This suggests that enhanced social support can mitigate the negative impact of sleep disturbances on anxiety and depression. Through quantitative analysis, we aim to reveal how varying levels of social support influence sleep quality’s predictive power for anxiety and depression. Our findings will provide targeted intervention strategies and scientific evidence to improve mental health outcomes among nursing interns.

## 2. Materials and methods

### 2.1 Participants

This study enrolled all nursing students interning at Quanzhou First Hospital in 2025. Inclusion criteria included: (1) Age ≥18 years; (2) Having signed an informed consent form to participate in the research. Exclusion criteria were: (1) Age under 18 years; (2) Personal or family history of mental health issues such as anxiety or depression; (3) Having received psychological counseling or therapy within the past year.

### 2.2 Measures

#### Basic Demographic Information Questionnaire

The basic demographic questionnaire, which we developed ourselves, includes two aspects: basic demographic information, such as age, gender, education and family situation; and behavioral habits, such as smoking, drinking and physical exercise.

#### Sleep Quality

The Pittsburgh Sleep Quality Index (PSQI) is a subjective assessment tool used in clinical and basic research to evaluate sleep quality, assessing the sleep quality of subjects over the past month ^[25]^. PSQI consists of 19 self-report items and 5 other-observer items, with the 19th self-report item and 5 other-observer items not contributing to scoring. The scored items can be combined into seven components: subjective sleep quality, sleep latency, sleep duration, habitual sleep efficiency, sleep disturbances, use of sleep medications, and daytime dysfunction. Each component is scored on a 0-3 scale, with cumulative scores summing to the total PSQI score (0-21 points), where higher scores indicate poorer sleep quality. A score >5 indicates poor sleep quality, while ≤5 indicates good sleep quality^[26-27]^. The Cronbach’s α-value of PSQI was reported as 0.83 in initial validation studies. Additionally, the questionnaire has been widely used in multiple regions^[28]^.

#### Social Support

This study adopted the Social Support Rate Scale (SSRS) to understand the characteristics of participants’ social support and its relationship with mental health and psychological disorders, which was used to measure individual social support status. The scale, developed by Xiao Shuiyuan in 1986, contains 10 self-rating items divided into three dimensions: subjective support (4 items), objective support (3 items), and availability support (3 items) ^[29]^. It is widely applied in China and demonstrates good validity and reliability^[30]^. The total score is calculated by summing the scores of the 10 items, with higher total scores indicating more adequate social support. Scores exceeding 44 indicate high-level social support, 23-44 points represent moderate levels, while scores below 23 indicate low levels^[31]^.

#### Anxiety

The Generalized Anxiety Disorder-7 (GAD-7) is a self-report questionnaire developed by Spitzer et al., designed to screen for generalized anxiety disorder and assess its severity based on symptoms reported over the past two weeks. This study employed this scale to evaluate participants’ anxiety levels^[32]^. As a self-report instrument, GAD-7 contains seven items with a four-point scoring system (0-3 points): 0 indicates no symptoms, 1 means occasional occurrences, 2 indicates more than half of days experience symptoms, and 3 means almost daily occurrences, with total scores ranging from 0 to 21 points. The primary outcome measure is the total sum score, which correlates positively with anxiety severity. Interpretation is based on validated cutoffs: scores of 0-4 are classified as minimal anxiety; 5-9, mild; 10-14, moderate; and scores of 15 or above indicate severe generalized anxiety^[33]^.

#### Depression

The Patient Health Questionnaire (PHQ-9) is a standardized psychological assessment tool administered through self-reporting^[34]^. It evaluates recent symptoms over the past two weeks to screen for and preliminarily diagnose depressive disorders and other common mental health conditions. This study utilized this questionnaire to measure participants’ depressive levels. As a self-report instrument, the PHQ-9 contains nine items with four response options: 0 (not at all), 1 (several days), 2 (more than half the days), and 3 (nearly every day). Total scores range from 0 to 27 points, where higher scores indicate more severe depression^[35-36]^. The scoring criteria are as follows: 0-4 points indicate no depression; 5-9 points for mild depression; 10-14 points for moderate depression; 15-19 points for moderate-to-severe depression; and 20-27 points suggesting possible major depressive disorder ^[37]^.

### 3.3 Procedures

We utilized the Wenjuanxing platform, one of the most widely used online survey platforms in China, to design an online questionnaire. From May to July 2025, during the pre-service training for nursing internships, we sent the survey link via WeChat and set a one-time completion requirement per participant. Before distributing the questionnaire, all participants were informed about the purpose of this study and the response method. An informed consent form was included at the beginning of the questionnaire, which participants needed to read and agree to. We also stated that the data collected from this survey would only be used for scientific research and would not disclose participants’ personal information. This study was conducted strictly in accordance with the Helsinki Declaration and has been formally approved by the Institutional Review Board (Approval No. Quan Yi Lun 2025K163). The entire questionnaire should take at least 5 minutes to complete, and samples with response times below this standard were excluded. Ultimately, we selected 396 participants as our final research subjects.

### 3.4 Data Analysis

We utilized SPSS version 26.0 for data cleaning, t-tests, ANOVA, normality tests, and correlation analyses. All statistical tests were performed using two-tailed methods with a significance level of

0.05. Additionally, we conducted data standardization for four key variables: sleep quality, social support, anxiety, and depression. Hierarchical regression analysis was employed to identify main effects and interactions between social support and sleep quality on anxiety and depression. Age, gender, and education level were treated as primary variables, with sleep quality as a secondary variable, social support as a tertiary variable, and their interaction term (SQ*SS) as a quaternary variable. A mediation model was constructed using AMOS 24.0 to examine how social support and sleep quality influence anxiety and depression in nursing interns. Interaction patterns were assessed through simple slope analysis to evaluate mediating effects. Results were visualized using online resources from www.jeremydawson.co.uk/slopes.htm^[38]^.

## 4.Results

### Demographics and descriptive analysis of the four key variables

This study collected 396 samples, with female participants constituting the majority (88.38%) and an average age of 20.56±0.69 years. Among the 355 nursing interns (89.65%), 253 (63.89%) held college degrees, while 253 (63.89%) were from rural areas. The vast majority of participants were not only children (85.86%) but also single-parent households (92.17%). Over 98.74% did not smoke and 99.50% abstained from alcohol (Table 1). As shown in Table 2, indicators such as sleep quality, social support, anxiety, and depression were influenced by demographic characteristics.

**Table 1.** Demographic information for participants.

| Variables | Model | N | % |
| --- | --- | --- | --- |
| Age |  | M=20.56, SD=0.69 |  |
| Gender | Male | 46 | 11.62 |
|  | Female | 350 | 88.38 |
| Education | College | 355 | 89.65 |
|  | Undergraduate | 41 | 10.35 |
| Family address | City | 143 | 36.11 |
|  | Village | 253 | 63.89 |
| One Child | Yes | 56 | 14.14 |
|  | No | 340 | 85.86 |
| Single-parent Family | Yes | 31 | 7.83 |
|  | No | 365 | 92.17 |
| Cigarette | Yes | 5 | 1.26 |
|  | No | 391 | 98.74 |
| Alcohol | Yes | 2 | 0.50 |
|  | No | 394 | 99.50 |
| Physical Activity | Never | 22 | 5.60 |
|  | Sometimes | 266 | 67.17 |
|  | 1-2 times a week | 79 | 19.95 |
|  | At least 3 times a week | 29 | 7.32 |

**Table 2.** Sleep quality, social support, anxiety and depression based On demographic characteristics.

| Variables | Model | N | Sleep Quality | Social Support | Anxiety | Depression |
| --- | --- | --- | --- | --- | --- | --- |
| Age<br>(years old) | <21 | 205 | 4.71±2.788 | 36.62±7.563 | 3.16±3.062 | 2.78±3.184 |
|  | ≥21 | 191 | 4.90±2.891 | 36.45±7.151 | 3.02±3.253 | 2.92±3.689 |
|  | *t* |  | -0.641 | 0.228 | 0.442 | -0.422 |
| Gender | Male | 46 | 4.72±2.680 | 33.89±7.770 | 3.52±3.371 | 2.83±3.573 |
|  | Female | 350 | 4.81±2.859 | 36.89±7.242 | 3.03±3.123 | 2.85±3.420 |
|  | *t* |  | -0.211 | -2.614 | 0.992 | -0.042 |
| Education | College | 355 | 4.60±2.715 | 36.90±7.414 | 2.92±3.020 | 2.49±2.983 |
|  | Undergraduate | 41 | 6.56±3.264 | 33.37±6.057 | 4.56±3.867 | 5.90±5.190 |
|  | *t* |  | -4.289*** | 2.944** | -3.196** | -6.310*** |
| Family Address | City | 143 | 4.82±3.001 | 35.9±7.662 | 2.86±3.082 | 2.31±3.148 |
|  | Village | 253 | 4.79±2.744 | 36.9±7.171 | 3.22±3.190 | 3.15±3.555 |
|  | *t* |  | 0.093 | -1.308 | -1.083 | -2.328* |
| One Child | Yes | 56 | 4.86±2.895 | 32.77±7.398 | 3.32±3.396 | 3.38±3.993 |
|  | No | 340 | 4.79±2.831 | 37.16±7.174 | 3.05±3.114 | 2.76±3.330 |
|  | *t* |  | 0.161 | -4.225*** | 0.597 | 1.245 |
| Single-parent Family | Yes | 31 | 4.97±3.371 | 34.23±8.127 | 2.65±3.421 | 2.52±3.295 |
|  | No | 365 | 4.79±2.791 | 36.73±7.267 | 3.13±3.131 | 2.87±3.447 |
|  | *t* |  | 0.342 | -1.828 | -0.815 | -0.557 |
| Cigarette | Yes | 5 | 7.40±2.302 | 30.60±6.731 | 4.60±2.302 | 5.20±2.683 |
|  | No | 391 | 4.77±2.829 | 36.61±7.343 | 3.07±3.159 | 2.82±3.434 |
|  | *t* |  | 2.071* | -1.821 | 1.079 | 1.546 |
| Alcohol | Yes | 2 | 4.50±2.121 | 34.00±0.000 | 6.00±2.828 | 4.00±2.828 |
|  | No | 394 | 4.80±2.841 | 36.55±7.375 | 3.07±3.151 | 2.84±3.438 |
|  | *t* |  | -0.150 | -0.489 | 1.311 | 0.476 |
| Physical Activity | Never | 22 | 7.14±3.808 | 30.86±5.321 | 5.82±4.856 | 6.0±6.597 |
|  | Sometimes | 266 | 4.67±2.611 | 36.52±6.986 | 2.98±2.949 | 2.62±2.914 |
|  | 1-2 times a week | 79 | 4.29±2.728 | 37.86±7.780 | 2.48±2.832 | 2.33±3.133 |
|  | At least 3 times a week | 29 | 5.59±3.397 | 37.38±9.006 | 3.69±3.242 | 3.97±3.941 |
|  | *F* |  | 7.055*** | 5.522*** | 7.267*** | 8.690*** |

Note: \*p < 0.05, \*\*p < 0.01, \*\*\*p < 0.001

### Correlations

Table 3 presents the correlations between sleep quality, social support, anxiety, and depression. Sleep quality showed a positive correlation with anxiety (r=0.552, P<0.01) and depression (r=0.653, P<0.01). However, the study revealed that social support exhibited negative correlations with sleep quality (r=-0.350, P<0.01), anxiety (r=-0.400, P<0.01), and depression (r=-0.390, P<0.01).

**Table 3.** Pearson correlations for sleep quality, social support, anxiety and depression.

| Variables | Mean±SD | $\alpha$ | 1 | 2 | 3 | 4 |
| --- | --- | --- | --- | --- | --- | --- |
| 1. Sleep Quality | 4.80±2.836 | 0.720 | 1.000 |  |  |  |
| 2. Social Support | 36.54±7.358 | 0.755 | -0.350** | 1.000 |  |  |
| 3. Anxiety | 3.09±3.153 | 0.885 | 0.552** | -0.400** | 1.000 |  |
| 4. Depression | 2.85±3.433 | 0.870 | 0.653** | -0.390** | 0.741** | 1.000 |

Note: \*\*p < 0.01

### The relationship between sleep quality, social support and anxiety

As shown in Table 4 and Figure 1, social support moderates the relationship between sleep quality and anxiety. In Model 2, sleep quality shows a significant correlation with anxiety, explaining 30.4% of the variance. In Model 3, sleep quality demonstrates a significant positive correlation with anxiety, while social support exhibits a significant negative correlation with anxiety, collectively explaining 34.7% of variance. In Model 4, sleep quality (β=0.449) shows a significant positive correlation with anxiety, whereas social support (β=-0.239) and their interaction term (β=-0.091) exhibit significant negative correlations with anxiety, increasing the variance explained by the interaction term by 0.7%. The degree of social support varies significantly across models, acting as a moderating factor in the relationship between sleep quality and anxiety. When social support is insufficient, sleep quality has a more pronounced impact on anxiety; conversely, when social support is adequate, this impact becomes relatively smaller (see Table 6 and Figure 2).

**Figure. 1.**
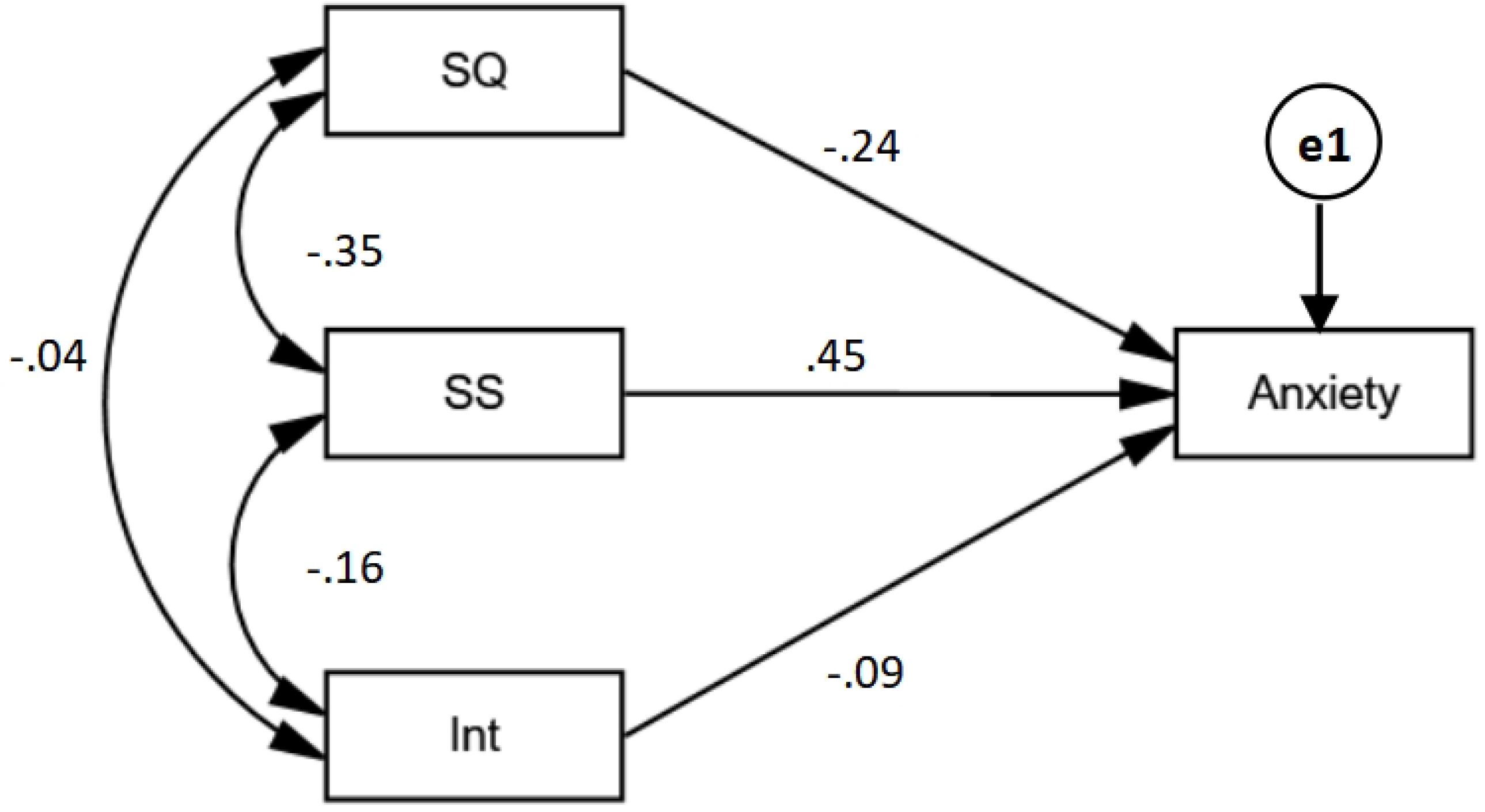

**Figure. 2.**
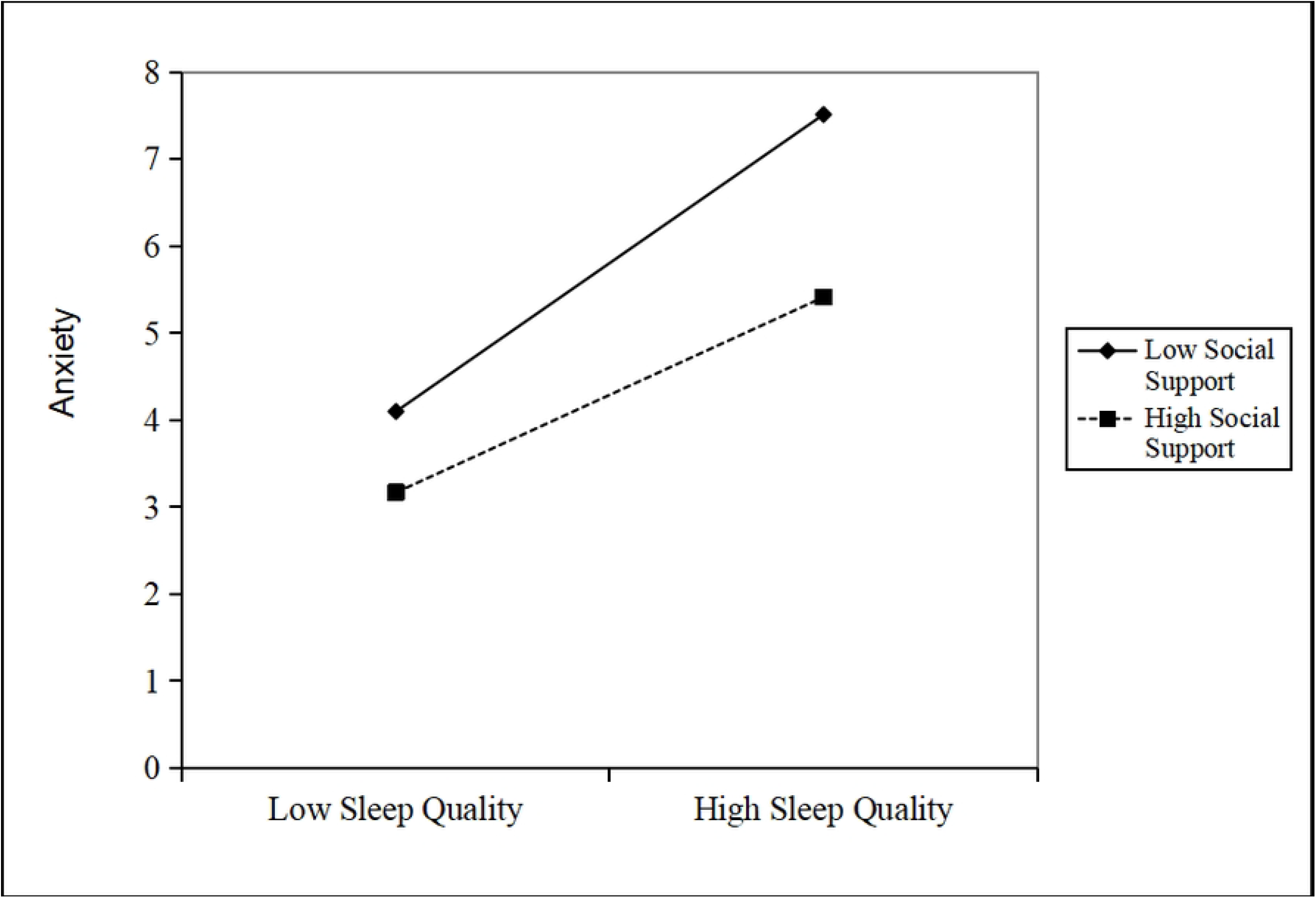

**Table 4.** Linear hierarchical regression to examine the interactive effect of sleep quality and social support on anxiety.

| | Predictor | $\beta$ | t | VIF | Adjust R square | $\Delta F$ | P |
| --- | --- | --- | --- | --- | --- | --- | --- |
| Model 1 | Age | -0.080 | -1.447 | 1.240 | 0.025 | 4.333 | 0.005** |
|  | Gender | -0.050 | -1.002 | 1.021 |  |  |  |
|  | Education | 0.190 | 3.463*** | 1.219 |  |  |  |
| Model 2 | Age | -0.042 | -0.889 | 1.246 | 0.304 | 158.472 | 0.000*** |
|  | Gender | -0.058 | -1.372 | 1.021 |  |  |  |
|  | Education | 0.059 | 1.237 | 1.281 |  |  |  |
|  | SQ | 0.542 | 12.589*** | 1.052 |  |  |  |
| Model 3 | Age | -0.031 | -0.674 | 1.248 | 0.347 | 26.919 | 0.000*** |
|  | Gender | -0.027 | -0.655 | 1.043 |  |  |  |
|  | Education | 0.039 | 0.835 | 1.290 |  |  |  |
|  | SQ | 0.465 | 10.529*** | 1.183 |  |  |  |
|  | SS | -0.228 | -5.188*** | 1.172 |  |  |  |
| Model 4 | Age | -0.020 | -0.450 | 1.261 | 0.354 | 4.846 | 0.028* |
|  | Gender | -0.025 | -0.617 | 1.043 |  |  |  |
|  | Education | 0.029 | 0.632 | 1.301 |  |  |  |
|  | SQ | 0.449 | 10.060*** | 1.217 |  |  |  |
|  | SS | -0.239 | -5.431*** | 1.188 |  |  |  |
|  | SQ*SS | -0.091 | -2.201* | 1.051 |  |  |  |

Note: \*p < 0.05, \*\*p < 0.01, \*\*\*p < 0.001, SQ: sleep quality, SS: social support

**Table 5.**
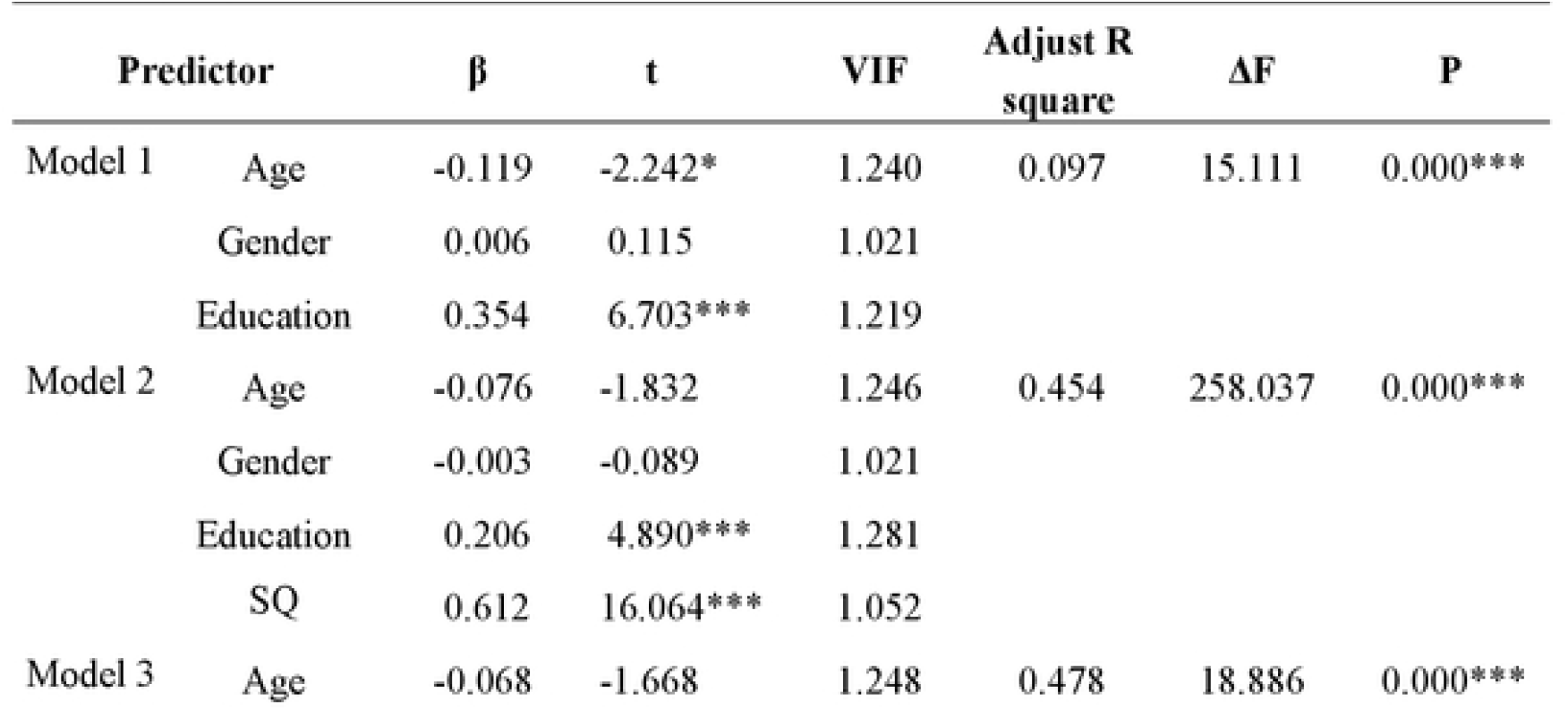
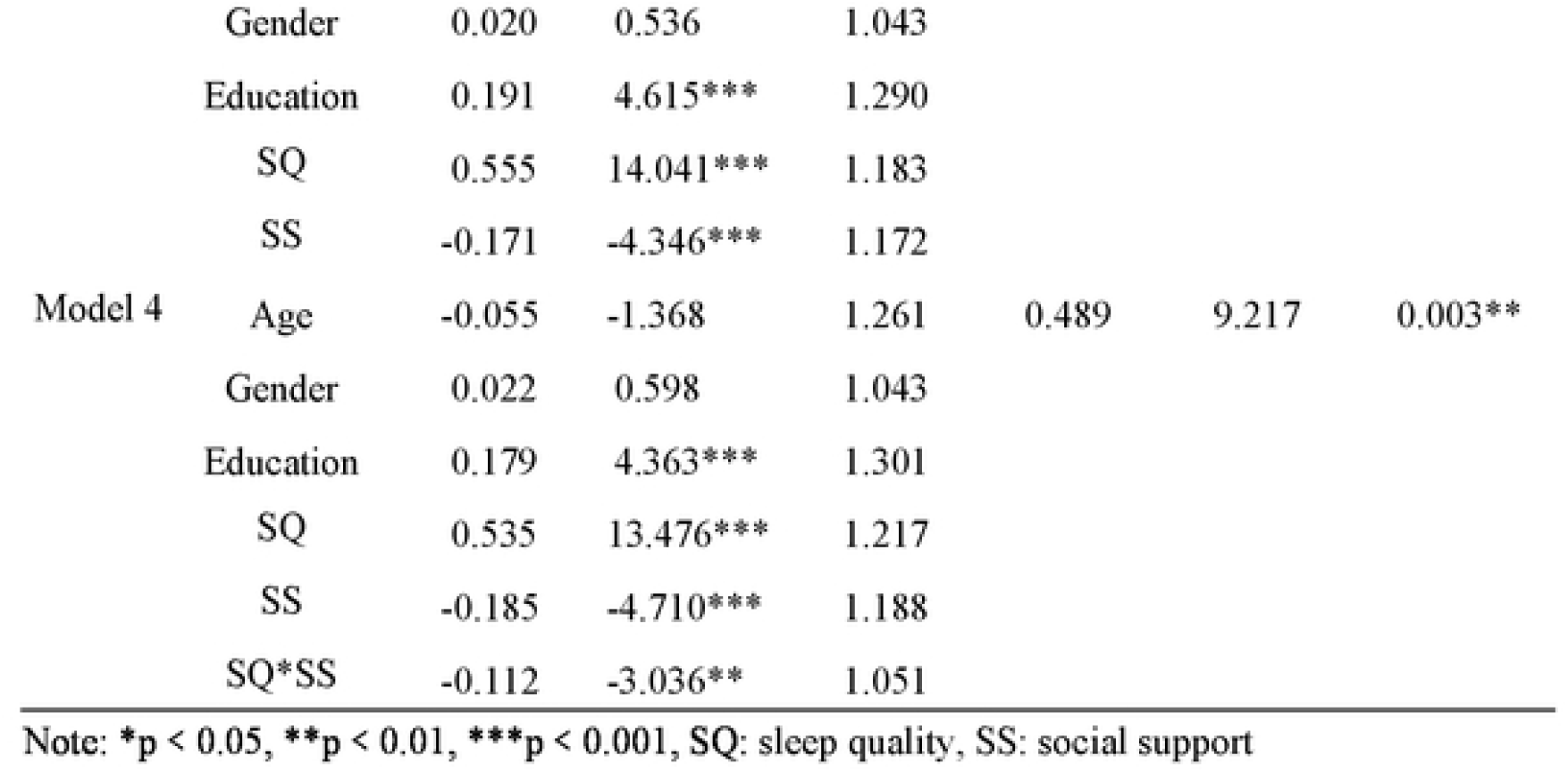
Linear hierarchical regression to examine the interactive effect of sleep quality and social support and depression.

| | Predictor | $\beta$ | t | VIF | Adjust R square | $\Delta F$ | P |
| --- | --- | --- | --- | --- | --- | --- | --- |
| Model 1 | Age | -0.119 | -2.242* | 1.240 | 0.097 | 15.111 | 0.000*** |
|  | Gender | 0.006 | 0.115 | 1.021 |  |  |  |
|  | Education | 0.354 | 6.703*** | 1.219 |  |  |  |
| Model 2 | Age | -0.076 | -1.832 | 1.246 | 0.454 | 258.037 | 0.000*** |
|  | Gender | -0.003 | -0.089 | 1.021 |  |  |  |
|  | Education | 0.206 | 4.890*** | 1.281 |  |  |  |
|  | SQ | 0.612 | 16.064*** | 1.052 |  |  |  |
| Model 3 | Age | -0.068 | -1.668 | 1.248 | 0.478 | 18.886 | 0.000*** |
|  | Gender | 0.020 | 0.536 | 1.043 |  |  |  |
|  | Education | 0.191 | 4.615*** | 1.290 |  |  |  |
|  | SQ | 0.555 | 14.041*** | 1.183 |  |  |  |
|  | SS | -0.171 | -4.346*** | 1.172 |  |  |  |
| Model 4 | Age | -0.055 | -1.368 | 1.261 | 0.489 | 9.217 | 0.003** |
|  | Gender | 0.022 | 0.598 | 1.043 |  |  |  |
|  | Education | 0.179 | 4.363*** | 1.301 |  |  |  |
|  | SQ | 0.535 | 13.476*** | 1.217 |  |  |  |
|  | SS | -0.185 | -4.710*** | 1.188 |  |  |  |
|  | SQ*SS | -0.112 | -3.036** | 1.051 |  |  |  |

Note: \* $p < 0.05$ , \*\* $p < 0.01$ , \*\*\* $p < 0.001$ , SQ: sleep quality, SS: social support

**Table 6.** Results of simple slope test of the effect of sleep quality on anxiety at different groups of social support.

| Group | Simple slope | *t* | *P* |
| --- | --- | --- | --- |
| Low SS | 0.602 | 4.340 | 0.000*** |
| High SS | 0.396 | 2.748 | 0.006** |

Note: \*\* $p < 0.01$ , \*\*\* $p < 0.001$ , SS: social support

### The relationship between sleep quality, social support, and depression

As shown in Table 5 and Figure 3, the relationship between sleep quality and depression is moderated by social support. The results indicate that in Model 2, sleep quality and education level show significant positive correlations with depression, explaining 45.4% of the variance. In Model 3, while both sleep quality and education level maintain significant positive correlations with depression, social support exhibits a significant negative correlation. These associations collectively account for 47.8% of the variance. Furthermore, in Model 4, sleep quality (β=0.535) and education level (β=0.179) demonstrate significant positive correlations with depression, whereas social support (β=-0.185) and their interaction term (β=-0.112) show significant negative correlations with depression. The interaction term contributes an additional 1.1% to variance explanation. Different levels of social support influence the relationship between sleep quality and depression differently. When social support is low, the impact of sleep quality on depression becomes significantly stronger; conversely, when social support is high, this impact relatively weakens (see Table 7 and Figure 4).

**Figure. 3.**
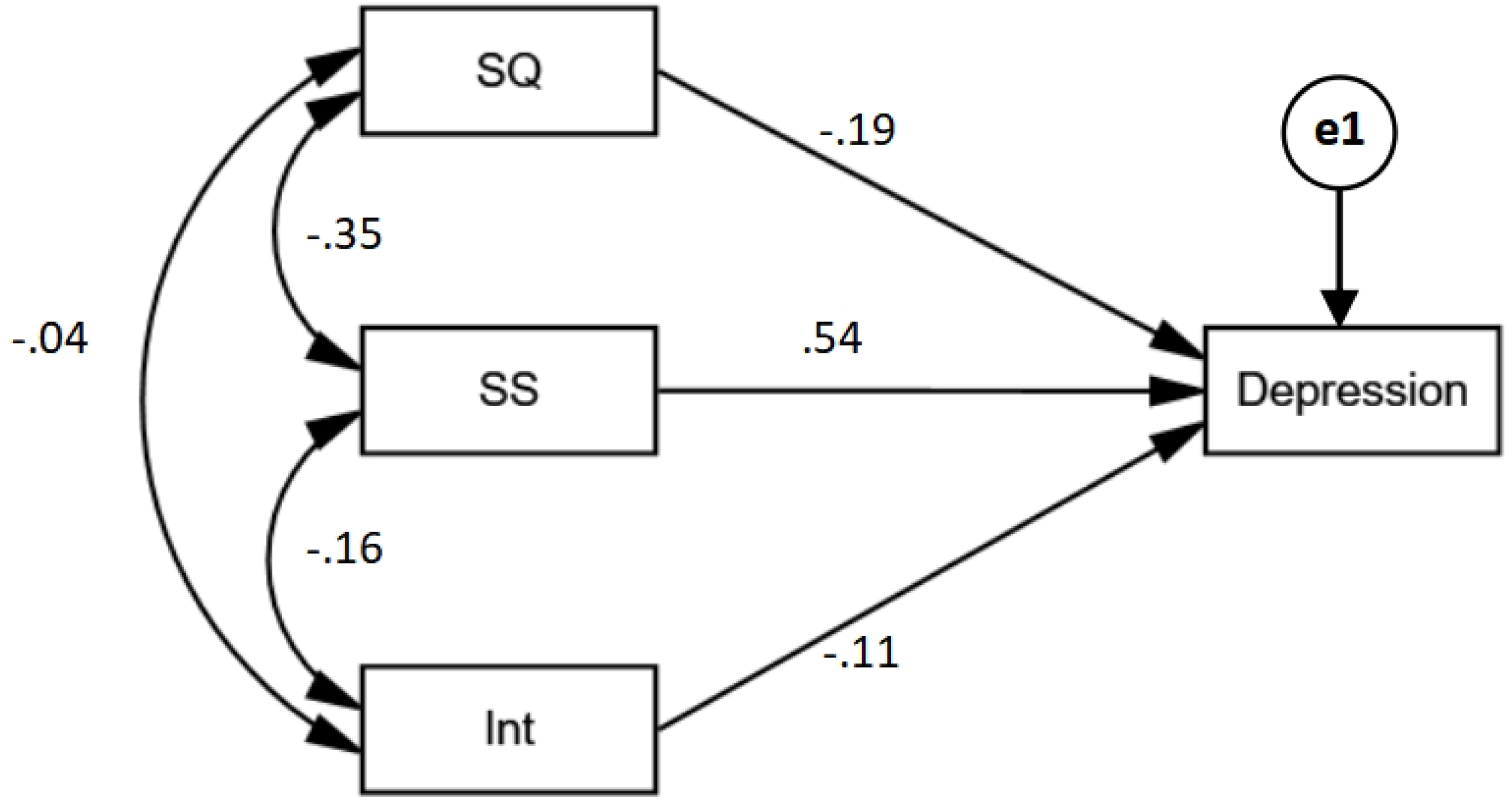

**Table 7.** Results of simple slope test of the effect of sleep quality on depression at different groups of social support.

| Group | Simple slope | *t* | *P* |
| --- | --- | --- | --- |
| Low SS | 0.779 | 5.811 | 0.000*** |
| High SS | 0.515 | 3.691 | 0.000*** |

Note: \*\*\* $p < 0.001$ , SS: social support

**Figure. 4.**
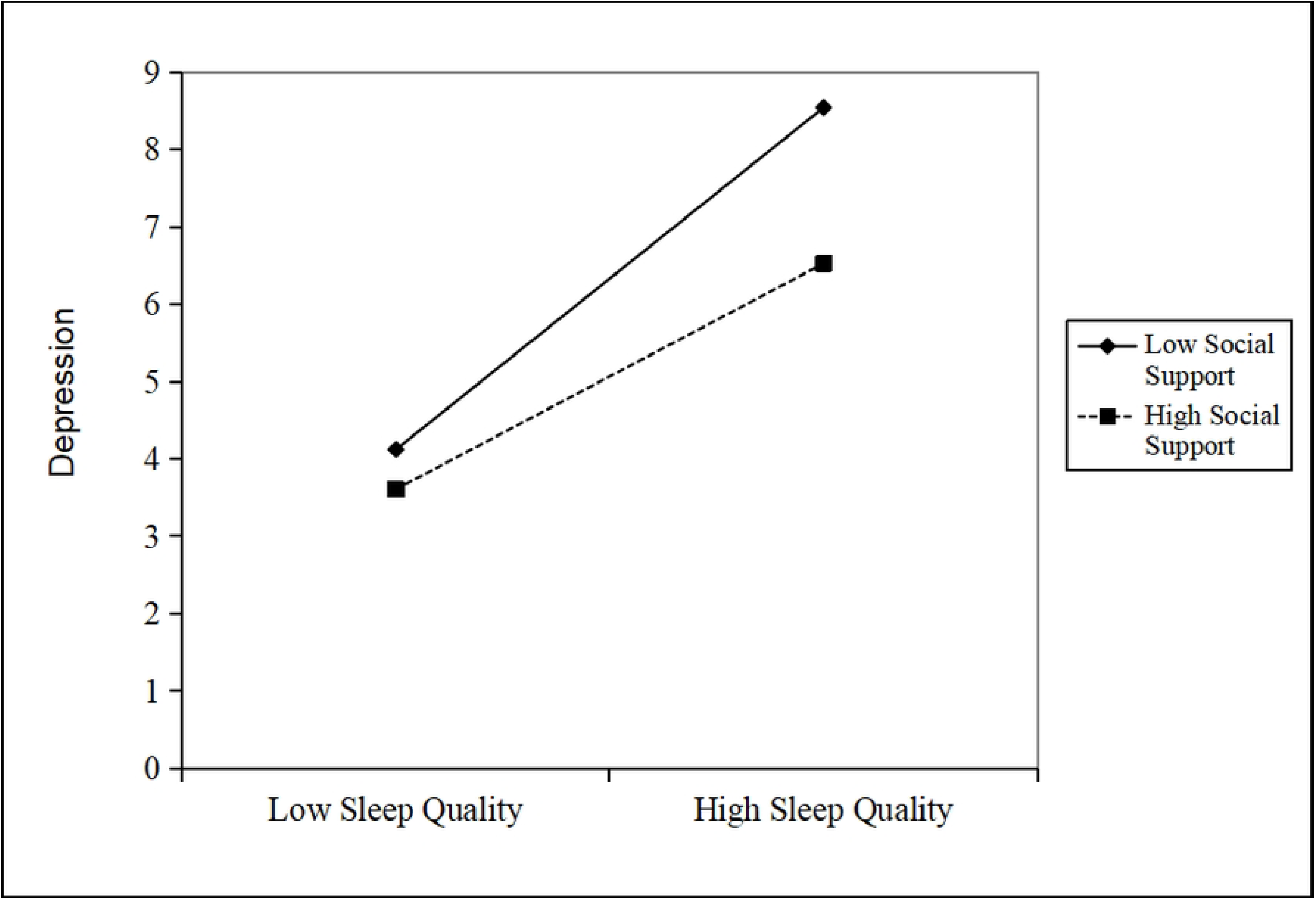

## 5.Discussion

### 5.1 The moderating role of social support

Through rigorous data collection and in-depth analysis, the study clearly reveals the significant moderating role of social support in the relationship between sleep quality and anxiety/depression. This finding holds important theoretical value and practical significance, providing additional evidence to deepen our understanding of the complex relationships among factors affecting mental health. Specifically, under conditions of low social support, the positive predictive effect of sleep quality on anxiety (β=0.602) and depression (β=0.779) is particularly prominent. This indicates that when nursing interns lack sufficient social support networks, the decline in sleep quality directly and strongly leads to the emergence and exacerbation of anxious and depressive emotions. As social support levels increase, the negative impact of sleep quality on anxiety (β= 0.396) and depression (β= 0.515) gradually diminishes. This suggests that strong social support creates an effective protective barrier, reducing the psychological impact of sleep issues on mental health. When nursing interns possess a broad and robust social support network, they can draw emotional comfort, informational guidance, and practical assistance from this network even when experiencing poor sleep quality. When facing sleep-related challenges, experienced mentors can provide professional advice to help adjust their daily routines. Family and friends offer care and encouragement, creating a sense of warmth and support that enhances stress resilience and alleviates psychological distress caused by sleep issues^[39-41]^.

From the perspective of mechanism, social support provides emotional comfort, which enables nursing interns to feel understood and accepted when facing stress, thus reducing psychological burden^[42-43]^. Information guidance provides them with methods and strategies to deal with problems and enhance their confidence in solving problems^[44]^. Practical help directly alleviates daily life difficulties, enabling them to handle various challenges more calmly. The combined effects of these aspects collectively enhance individuals’ stress resilience and effectively alleviate psychological distress caused by sleep issues^[45]^.

### 5.2 The relationship between sleep quality and mental health

The study once again provides conclusive evidence confirming a significant positive correlation between sleep quality and anxiety (r=0.552, P<0.01) and depression (r=0.653, P<0.01), with this relationship being particularly pronounced among nursing interns. Due to the unique demands of their profession, nursing interns face compounded challenges including shift work schedules, intensive clinical responsibilities, and academic pressures, leading to widespread issues of declining sleep quality^[46-48]^.

Rotating work disrupts the normal biological clock of the human body, resulting in sleep rhythm disorder^[49]^. Nursing interns may need to frequently switch between day and night work patterns, which makes it difficult for their bodies to adapt to regular sleep time, thus affecting sleep quality^[50]^. High intensity clinical tasks require them to keep a high degree of concentration and alertness at all times, and their mental state is under tension for a long time. Even after work, it is difficult to relax quickly, which further affects the sleep and depth of sleep^[51]^. At the same time, academic pressure should not be underestimated. Nursing interns need to complete clinical work while taking into account theoretical learning and examinations, which undoubtedly increases their psychological burden and further reduces the time for rest and sleep^[52-52]^.

The decline in sleep quality can trigger a range of physical and psychological issues, which may further increase susceptibility to mood disorders. Physiologically, insufficient sleep disrupts normal brain function by causing neurotransmitter imbalances that impair emotional regulation^[54-56]^. From a psychological perspective, long-term poor sleep will make nursing interns feel tired, helpless and depressed, and prone to anxiety and depression^[57-58]^. Ensuring nursing interns maintain quality sleep is essential for building a solid foundation for their mental health. Healthcare institutions and educational departments should fully recognize this need and implement effective measures to improve sleep conditions, such as scheduling work hours rationally, providing comfortable rest environments, and conducting sleep health education programs.

### 5.3 Theoretical support: stress buffer model

The findings of this study are highly consistent with the stress-buffering model of social support, providing strong evidence for its applicability in the context of nursing interns. The stress-buffering model posits that social support can mitigate the negative impact of stressors on individual mental health through multiple pathways^[59]^. Specifically, social support can reduce individuals’ perception of stress. When nursing interns face stressors like sleep issues, support from peers, mentors, and family helps them feel they’re not alone, thereby alleviating fear and anxiety about pressure^[60]^. At the same time, social support can also promote the formation of adaptive coping strategies^[61]^. When facing stress from sleep-related issues, nursing interns with strong social support are more likely to adopt proactive and effective coping strategies such as seeking professional help or adjusting their lifestyle. Experienced mentors can provide practical advice and guidance to help them find solutions. Meanwhile, family and friends can offer daily supervision and encouragement, fostering the development of healthy sleep habits in interns. In addition, social support can enhance individual psychological resilience^[62-63]^. Psychological resilience refers to an individual’s ability to recover and adapt quickly when facing stress and adversity. Support from various sectors of society can empower nursing interns to become stronger and more confident in overcoming challenges related to sleep issues^[64-66]^.

In conclusion, social support plays a crucial mediating role in the relationship between sleep quality and mental health among nursing interns. Given the close connection between sleep quality and psychological well-being, the stress buffer model provides robust theoretical support for understanding this relationship. To promote the mental health of nursing interns, medical institutions, educational departments, and society at large should collaborate to provide comprehensive social support, improve sleep quality, and create a conducive environment for their growth and development^[67-68]^.

## 6. Limitations

Several limitations should be acknowledged. First, this study employed a cross-sectional design, which precludes causal inferences regarding the relationships among sleep quality, social support, and mental health. Longitudinal or experimental designs are needed to establish temporal precedence and causality. Second, all measures were self-reported, which may introduce response bias such as social desirability or recall inaccuracy. Future studies could benefit from incorporating objective sleep assessments (e.g., actigraphy) and multi-informant reports of social support. Third, the sample was recruited from a single hospital in Quanzhou, which may limit the generalizability of findings to nursing interns in other regions or healthcare systems. Multi-site studies across diverse cultural and institutional contexts are warranted. Fourth, although we controlled for basic demographic variables, other potential confounders such as personality traits, coping styles, or clinical workload were not assessed. These factors may influence both sleep and mental health outcomes and should be considered in future research. Lastly, the exclusion of individuals with pre-existing mental health conditions or prior psychological treatment may have resulted in an underestimation of the true prevalence of anxiety and depression in this population, potentially limiting the external validity of our findings.

## 7. Conclusions

This study demonstrates that sleep quality is a significant predictor of anxiety and depression among Chinese nursing interns. Social support moderates this relationship, with higher levels of support attenuating the negative effects of poor sleep. These findings highlight the dual importance of addressing sleep disturbances and enhancing social support systems in clinical training environments. Health policymakers, educational institutions, and hospital administrators should collaborate to develop comprehensive mental health promotion strategies that include sleep management programs and social support initiatives. Such efforts could improve the well-being of nursing interns, ultimately contributing to a more resilient healthcare workforce.

## Data Availability

All relevant data are within the manuscript and its Supporting Information files.

## Additional information and declarations

## 1.Glossary

PSQI: Pittsburgh Sleep Quality Index
SSRS: Social Support Rate Scale
GAD-7: the 7-item Generalized Anxiety Disorder Questionnaire
PHQ-9: Patient Health Questionnaire-9;
ANOVA: Analysis of Variance

## 3. Author Contributions

Yanyan Zhao: Project administration, Investigation, Data curation, Formal analysis, Writing-original draft, Writing-review & editing.

Fuzhi Liu: Data curation, Formal analysis, Methodology, Writing-original draft, Writing-review & editing.

Lanlan Chen: Resources, Conceptualization. Xiuli Li: Investigation.

Zhuote Tu: Software, Visualization.

Biyu Wu: Supervision, Writing-review & editing.

All authors contributed to the article and approved the submitted version.

## 5. Institutional Review Board Statement

The study was approved by the institution review board of Quanzhou First Hospital (No. Quan Yi

Lun 2025K163). All subjects provided their written informed consent to participate in this study.

## 6.Acknowledgments

We thank the participants in the study and acknowledge the reviewers and editors for viewing our manuscript.

## 7. Data Availability Statement

The original contributions in the study are included in the article, and further inquiries can be made directly to the corresponding author.

